# Surveillance of colistin-resistance and *mcr genes in* multi-drug resistant Enterobacteriaceae

**DOI:** 10.1101/2021.05.21.21255172

**Authors:** Yasuhide Kawamoto, Norihito Kaku, Norihiko Akamatsu, Kei Sakamoto, Kosuke Kosai, Yoshitomo Morinaga, Norio Ohmagari, Koichi Izumikawa, Yoshihiro Yamamoto, Hiroshige Mikamo, Mitsuo Kaku, Kazunori Oishi, Katsunori Yanagihara

**Author notes:** **Address for correspondence**: Norihito Kaku, Department of Laboratory Medicine, Nagasaki University Graduate School of Biomedical Sciences, 1-7-1 Sakamoto, Nagasaki, Nagasaki 852-8501, Japan.

## Abstract

**Background:** The plasmid-mediated bacterial colistin-resistant (*mcr*) gene is a global concern in clinical health care. This study aimed to clarify the prevalence of colistin resistance through nine *mcr* genes in ESBL-producing and CRE isolated Enterobacteriaceae in Japan.

**Methods:** We collected strains from August 2016 to March 2017 from five tertiary hospitals. MICs were measured using the microdilution method. PCR was performed to detect *mcr-1* to *mcr-9* genes in all strains. Additionally, we performed whole-genome sequencing of the *mcr* gene-positive strain.

**Results:** The rate of colistin resistance was 7.7%. The *mcr-5* and *mcr-9* gene were detected in one ESBL-producing *E. coli* strain (0.37%) and three CRE strains (1.1%), respectively. Since the ESBL-producing *E. coli* strain was the first clinical strain with *mcr-5* in Japan, whole-genome sequencing analysis was performed for the strain. The sequenece type of the *mcr-5* positive strain was ST1642 and it carried two distinct plasmids, ESBL gene-carrying pN-ES-6-1 and *mcr-5*.*1*-carrying pN-ES-6-2.

**Conclusions:** We showed that the frequency of colistin resistance and *mcr*-positive strains is not high in Japan. Since the MIC for colistin was low in the *mcr-5*.*1* and *mcr-9* gene-positive strain, continuous monitoring of *mcr* genes is necessary.

## Introduction

The emergence and spread of antimicrobial resistance are a global concern. In Japan, a previous study reported that ESBL-producing *Escherichia coli* (*E. coli*) and *Klebsiella pneumoniae* (*K. pneumoniae*) strains are spreading, accounting for 23.0% of *E. coli* and 10.7% of *K. pneumoniae* infections [1]. Because of the distribution of fluoroquinolone resistant in ESBL-producing *E. coli*, the clinical use of carbapenem is increasing [2, 3]. However, in recent years, carbapenem-resistant Enterobacteriaceae (CRE) have become a serious problem worldwide. Some CREs have multidrug resistance against fluoroquinolone as well as beta-lactam [4, 5]. Therefore, colistin, which belongs to the family of polymyxins and has broad-spectrum activity against gram-negative bacteria, is an important antibiotic in the treatment of CRE and ESBL.

On the other hand, several colistin resistance mechanisms have been reported. The major colistin resistance mechanisms are as follows: alteration of the LPS moiety resulting in a reduced net negative charge of LPS, increased drug efflux, overexpression of outer membrane protein (OprH), and the formation of capsules (*siaD, ompA, cps*) [6, 7]. Since all of these resistance mechanisms are intrinsic, mutational, and adaptive, colistin resistance is unlikely to spread from cell to cell through delivery of plasmids like ESBL and carbapenemase-producing Enterobacteriaceae (CPE). However, in 2016, the first plasmid-mediated colistin resistance (*mcr*) gene was identified in animals in China [8]. Thereafter, *mcr*-positive Enterobacteriaceae have been identified in healthy people and patients all over the world [9]. In Japan, although there have been some reports on *mcr*-positive *E. coli* in animals and food sources [10-12], there are few studies in humans [13]. In addition, the percentage of colistin resistance in Japan remains unknown because the MIC of colistin in Enterobacteriaceae including ESBL and CRE has not been evaluated.

The purpose of this study was to clarify the prevalence of colistin resistance and nine *mcr* genes in ESBL-producing Enterobacteriaceae and CRE isolated from patients in tertiary hospitals in Japan. Additionally, we performed whole-genome sequencing of the *mcr* gene-positive strain.

## Materials and methods

### Strains

A total of 273 different clinical ESBL-producing Enterobacteriaceae and CRE isolates between August 2016 and March 2017 were collected from five tertiary hospitals representing the Western, Eastern, and Central regions of Japan. Only one isolate per patient was included in this study. *Proteus* spp. and *Providencia rettgeri* were excluded from analysis, because they are intrinsically resistant to colistin. A total of 180 ESBL-producing strains and 93 CRE strains were collected during the study period. The ESBL-producing strains contained E. coli (81.7%), Klebsiella pneumoniae (15.6%) and Klebsiella oxytoca (2.8%). The CRE strains contained Klebsiella aerogenes (45.2%), Enterobacter cloacae complex (38.7%), Klebsiella pneumoniae (11.8%), E. coli (2.2%) and Citrobacter spp. (2.2%). The strains were stored in a Microbank tube and placed at -80°C.

### Analysis of strains

Bacteria were identified using MALDI-TOF MS (Bruker Daltonics GmbH, Bremen, Germany). ESBL production was detected using the BD Phoenix system NMIC-207 panel (Becton Dickinson, Holdrege, USA) according to the manufacturer’s instructions. Carbapenemase genes were evaluated by Xpert Carba-R assay (Cepheid, Sunnyvale, USA) according to the manufacturer’s instructions. MICs were measured using the microdilution method. Susceptibility was determined according to the CLSI M100-S25 except for colistin, which was interpreted according to the EUCAST version 9.0 (MIC for susceptible, ≤ 2 mg/L; MIC for resistant, >2 mg/L).

### Analysis of mcr genes

PCR was performed to detect *mcr-1* to *mcr-9* genes in all strains. DNA was extracted using the boiling method with minor modifications [14]. PCR amplification about *mcr-1* to *mcr-5* was performed under the following conditions: 15 min at 94°C, 25 cycles of 30 s at 94°C, 30 s at 58°C, 60 s at 72°C, and 10 min at 72°C for the final extension [15]. PCR amplification about *mcr-6* to *mcr-9* was performed under the following conditions: 3 min at 95°C, 30 cycles of 30 s at 95°C, 30 s at 55°C, 60 s at 72°C, and 10 min at 72°C for the final extension[16].

### Whole-genome sequencing

DNA was extracted using the Quick-DNA™ Fecal/Soil Microbe Miniprep kit according to the manufacturer’s instructions (Zymo Research, CA, USA). Whole-genome sequencing was performed using NextSeq 500 (Illumina Inc. San Diego CA USA) and GridION X5 (Oxford Nanopore Technologies, Oxford, UK). The *de novo* hybrid assembly of both short-reads (NextSeq 500) and long-reads (GridION X5) was performed using Unicycler v0.4.7 under conservative conditions. CheckM v1.0.12 was used to assess the quality of assembled genomes. The allele sequences and sequence types (STs) were determined according to the *E. coli* database (http://mlst.warwick.ac.uk/mlst/dbs/Ecoli). In the plasmid analysis, Prokka v1.13 was used for genome annotation. Antimicrobial resistance genes were identified using ResFinder v3.2 (https://cge.cbs.dtu.dk/services/ResFinder/). The bacterial insertion sequence was detected using the IS Finder database (https://isfinder.biotoul.fr). PlasmidFinder v2.0.1 was used to determine plasmid incompatibility (Inc) groups. Importing Prokka’s annotation result and drawing the plasmid map with Snap Gene v4.3.10 GSL Biotech.

### Phylogenetic analysis and genetic environment

The *mcr-5-*carrying plasmid sequencing from *Salmonella enterica* (Gene accession no. NC_003277.2), *Salmonella enterica* (Gene accession no. LC488708.01), *Escherichia coli* (Gene accession no. BENI01000099.1), pN-ES-6-2 *Escherichia coli* and *Salmonella enterica* (Gene accession no. KY807921.1) were downloaded from GenBank database. Phylogenetic tree and genetic environment were constructed using CLC Genomics Workbench version 21.0.3.

### Data availability

Raw data were generated at Nagasaki University Hospital. Derived data supporting the findings of this study are available from the corresponding author upon request.

### Ethics

This study was approved by the Ethics Committee of Nagasaki University Hospital (16072509). Data regarding clinical ESBL-producing Enterobacteriaceae and CRE isolates were anonymized and individually numbered when they were collected from the hospitals.

## Results

### Colistin Susceptibility

The rate of colistin resistance in all strains was 11.4% (Table 1). The rate of colistin resistance in the CRE strains was higher than that in the ESBL-producing strains (7.7% versus 18.9%, Table 1). The MIC of colistin in both ESBL-producing strains and CRE formed a bimodal distribution (Fig. 1A and B). The *Enterobacter cloacae* complex had the highest rate of colistin resistance (50.0%) (Table 1). The rate of colistin resistance was not significantly different based on site of infection or region where the strain was isolated (Table S1). In the CRE strains, carbapenemase-producing Enterobacteriaceae (CPE) had a higher rate of colistin resistance (40.0%) than non-CPE strains (16.6%) (Fig. 1C).

**Table 1.** Colistin resistance in ESBL-producing and CRE strains

| Organisms | Colistin-resistance |  | Carbapenemase genes<br>(number) | *mcr* genes<br>(number) |
| --- | --- | --- | --- | --- |
|  | n | (%) |  |  |
| ESBL-producing strains (180) | 2 | (1.1 %) |  |  |
| *Escherichia coli* (147) | 1 | (0.68 %) | N.D. | *mcr-5* (1) |
| *Klebsiella pneumoniae* (28) | 1 | (3.6 %) | N.D. | N.D. |
| *Klebsiella oxytoca* (5) | 0 | (0.0%) | N.D. | N.D. |
| CRE strains (93) | 19 | (20.4%) |  |  |
| *Citrobacter* spp. (2) | 0 | (0.0%) | N.D. | N.D. |
| *Escherichia coli* (2) | 0 | (0.0%) | N.D. | N.D. |
| *Enterobacter cloacae* complex (36) | 18 | (50.0%) | IMP-1 group (6) | *mcr-9* (3) |
| *Klebsiella aerogenes* (42) | 1 | (2.4%) | IMP-1 group (1) | N.D. |
| *Klebsiella pneumoniae* (11) | 0 | (0.0%) | IMP-1 group (7),<br>NDM and OXA-48 (1) | N.D. |
| Total (273) | 21 | (7.7%) |  |  |

N.D., not detected

**Figure 1.**
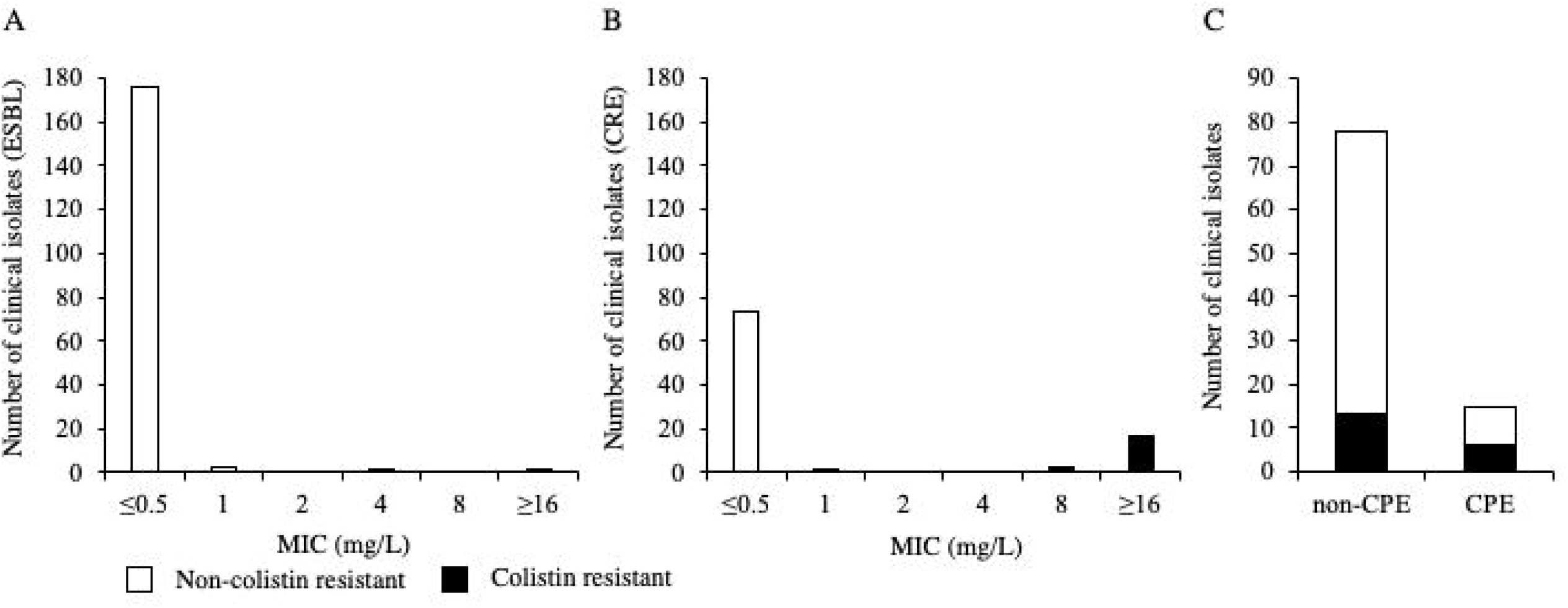
The Distribution of colistin minimum inhibitory concentration (MIC) in ESBL producing strains and CRE strains, Japan, August 2016-March 2017 (A). B) The comparison of colistin resistance rates between non-CPE and CPE. The MICs of colistin were measured using microdilution method. Blue indicates no-colistin resistant; red, colistin resistant.

**Figure 2.**
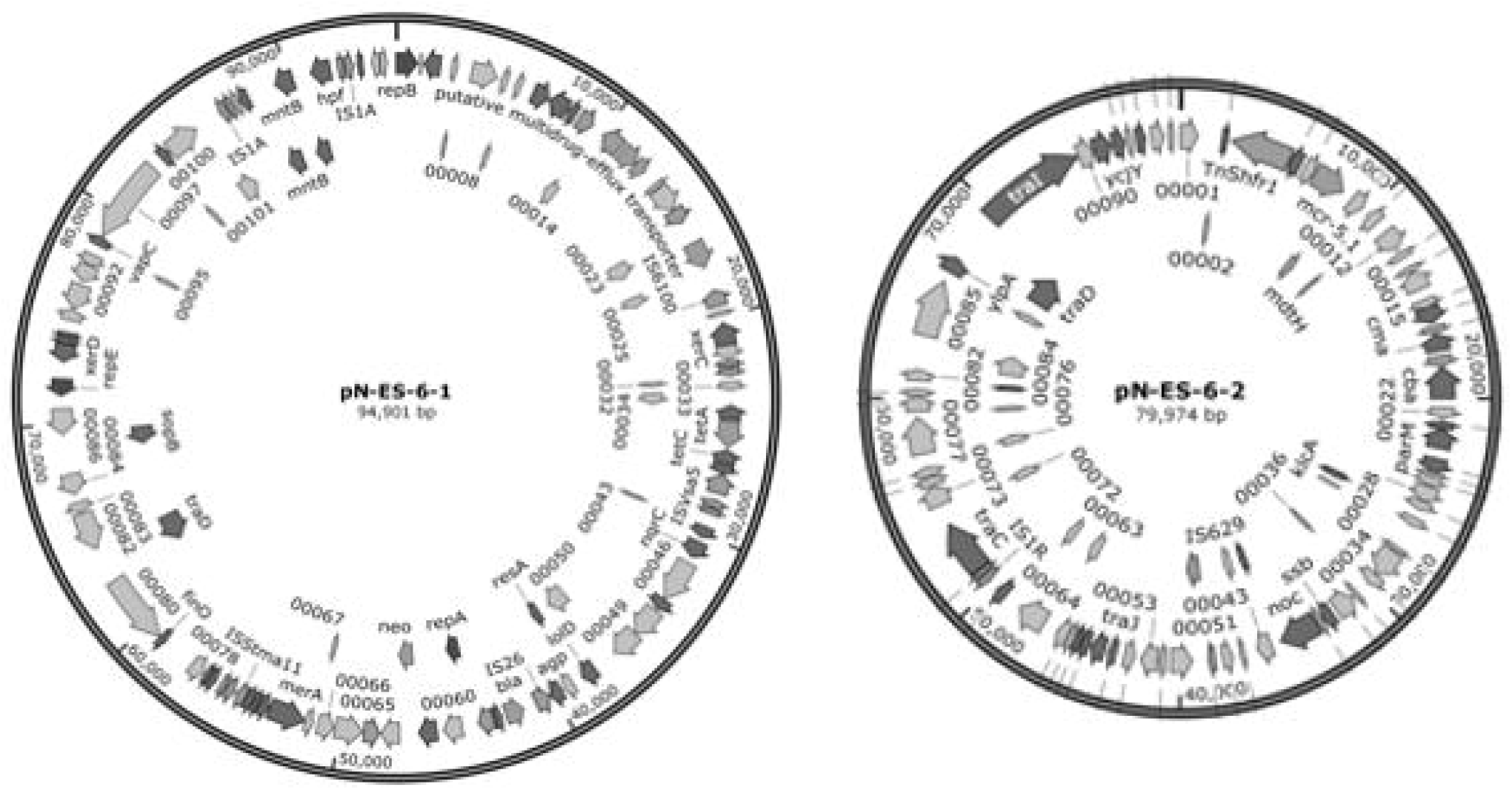
Structure of plasmid pN-ES-6-1 and pN-ES-6-2 from *Escherichia coli*. Yellow green indicates antimicrobial resistant; gray, hypothetical protein; red, insertion sequence; green, plasmid replication; purple, other protein.

### Detection of mcr genes

The 273 strains (ESBL and CRE) were screened using PCR for the presence of the nine *mcr* genes, *mcr-1* to *mcr-9*. The *mcr-5* gene was detected in only one ESBL-producing *E. coli* strain (0.37%), and the *mcr-9* gene was detected in three CRE *Enterobacter cloacae* complex strains (1.1%). The two of three *mcr-9* positive strains were isolated in the same hospital, but the ward and the department were different. All the *mcr* positive strains indicted very low MIC for colistin (Table S2).

### Whole-genome sequencing of mcr-5 gene-positive strain

Since there were no reports of *mcr-5* positive ESBL-producing *E. coli* infecting humans in Japan, we conducted a detailed analysis by whole-genome sequencing. The draft genome of *mcr-5* gene-positive *E. coli* strain (DDBJ accession no. DRA010253) comprised 5,027,748 bp with an overall GC content of 50.76%. The genome consisted of 22 rRNA operons, 80 tRNA genes, and 4686 protein-coding genes (CDSs). The *mcr-5* gene-positive *E. coli* was classified as ST1642 and *fimH* subtype 31. The isolate harbored two distinct plasmids, pN-ES-6-1 (DDBJ accession no. LC553463, 94901 bp), and pN-ES-6-2 (DDBJ accession no. LC553464, 79974 bp). pN-ES6-1 had various resistance genes, including *mph*(A), *dfrA17, bla*_TEM-1B_, *sul2, tet*(B), *aac(3)-lld, aac(3’’)-lb*, and *aph(6)-ld*, whereas pN-ES-6-2 had only one resistance gene (*mcr-5*.*1*) (Table 2). The Plasmid Inc. groups and transposons of pN-ES-6-2 were IncFII and TnShfr1 (Tn3-family), respectively (Table 2). Phylogenetic analysis of *mcr-5* carrying plasmid revealed that pN-ES-6-2 showed similarities to *Escherichia coli* (Gene accession no. BENI01000099.1) isolated in Japan, but less to the first reported *mcr-5* carrying plasmid (Gene accession no. KY807921.1, *Salmonella enterica*, Germany) (Figure 3 A). pN-ES-6-2 lacked some major facilitator superfamily (MFS) gene in comparison with other *mcr-5* carrying plasmids (Figure 3 B).

**Table 2.**
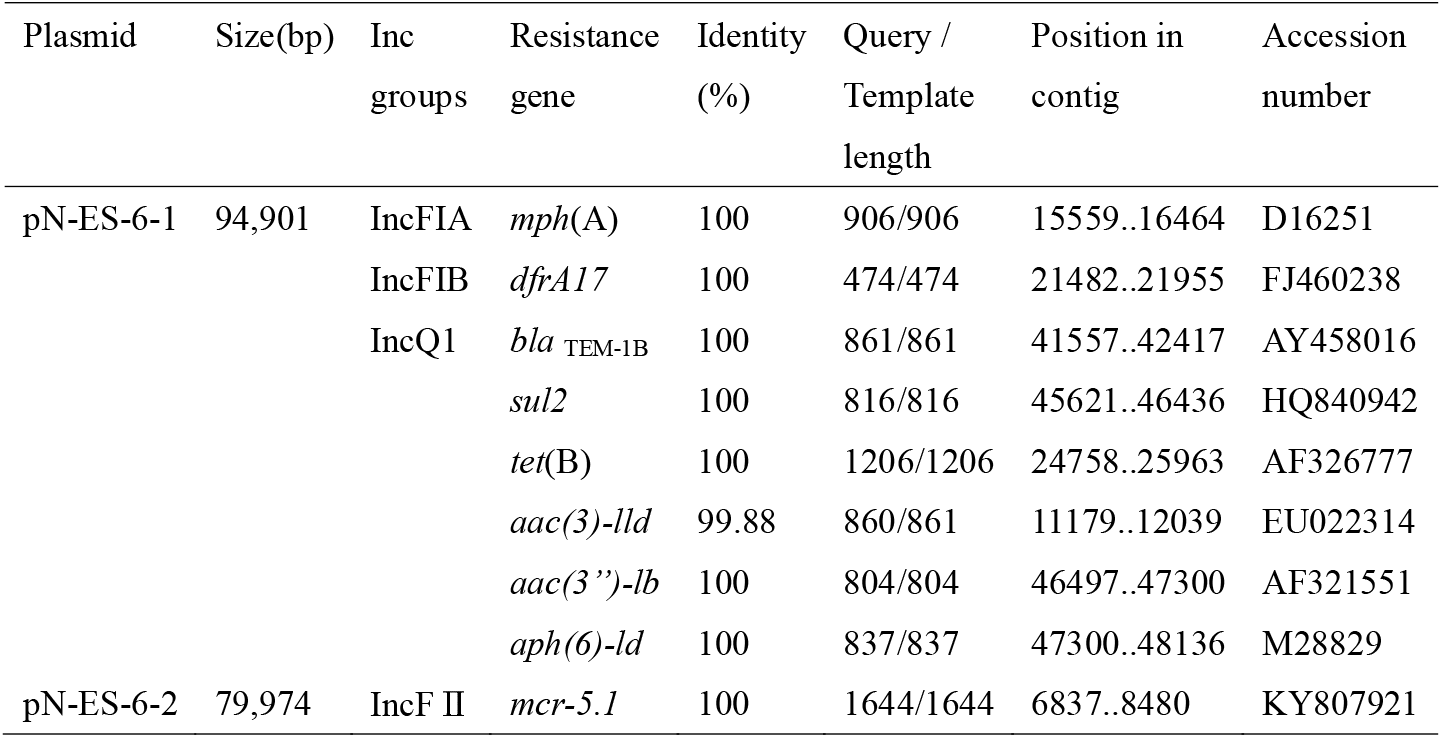
Inc groups and antimicrobial resistance genes of *mcr-5* gene positive strain

**Figure 3.**
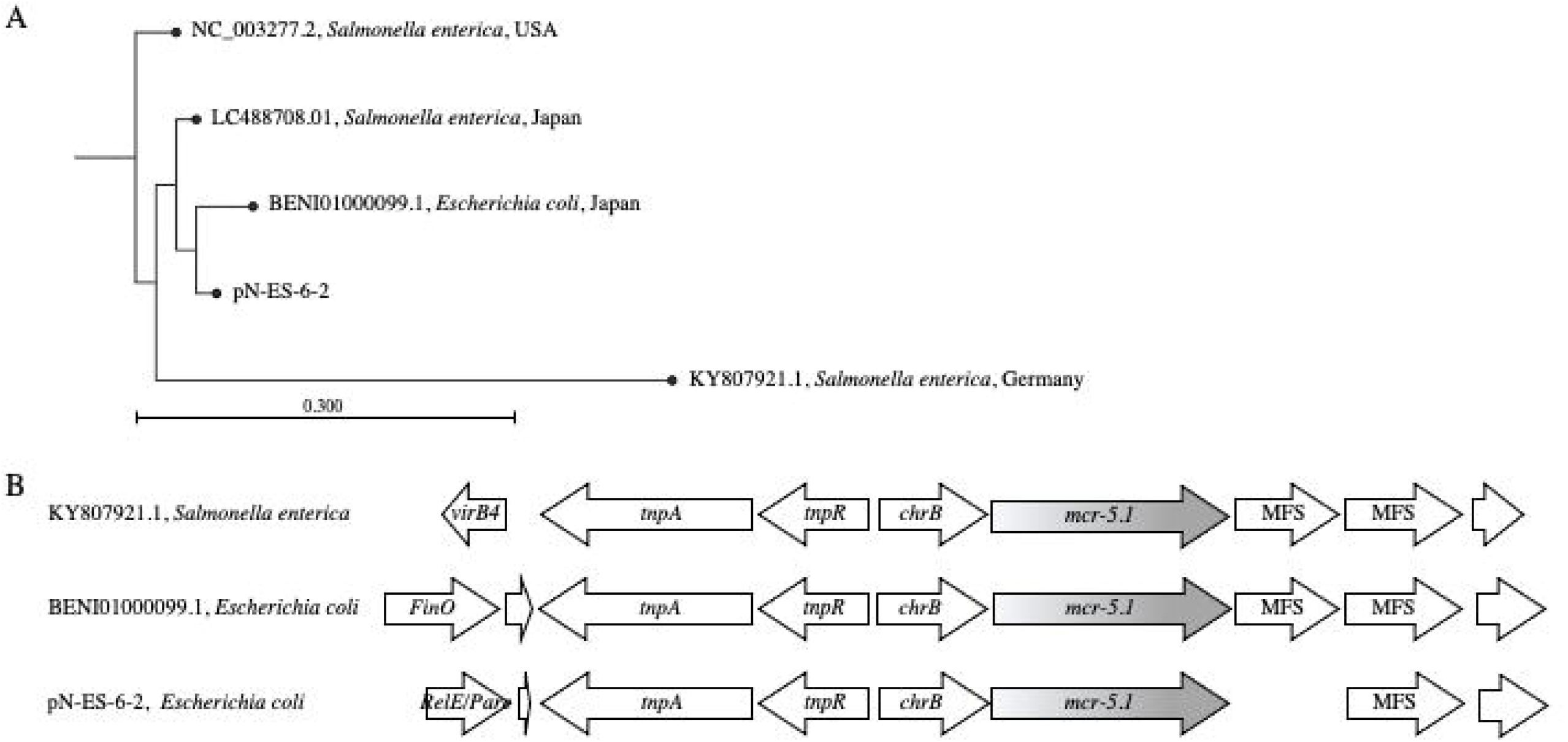
Comparison of *mcr-5-carrying* plasmid. A) Phylogenetic tree of *mcr-5-carrying* plasmid from *Salmonella enterica* (Gene accession no. NC_003277.2), *Salmonella enterica* (Gene accession no. LC488708.01), *Escherichia coli* (Gene accession no. BENI01000099.1), pN-ES-6-2 *Escherichia coli* and *Salmonella enterica* (Gene accession no. KY807921.1). Arrow represent coding sequences (gray arrows, *mcr-5*) and indicated direction of transcription. B) The genetic environment of *mcr-5* from *Salmonella enterica* (Gene accession no. KY807921.1), *Escherichia coli* (Gene accession no. BENI01000099.1) and pN-ES-6-2 *Escherichia coli*.

## Discussion

In this study, we clarified the prevalence of colistin resistance in ESBL-producing Enterobacteriaceae and CRE. The rates of colistin resistance were 0.7% for *E. coli*, 2.6% for *K. pneumonia*, and 50.0% for *Enterobacter cloacae* complex. Global surveillance in 2015 reported that the resistance rate of colistin was 0.3% in *E. coli*, 2.4% in *K. pneumonia*, and 39.1% in *Enterobacter asburiae* [17]. The Surveillance of Multicenter Antimicrobial Resistance in Taiwan reported that the resistance rate of colistin was 0.3% in *E. coli* and 2.4% in *K. pneumonia* [18]. These results are consistent with our study. Accordingly, in Japan, it is considered that colistin-resistant bacteria have not become widespread. However, in the CRE strains, the rate of colistin resistance in CPE strains was higher than that in non-CPE strains. A previous study reported a strong association between the presence of carbapenemase and increased resistance to colistin in Enterobacteriaceae strains [17]. In addition, other investigators have reported clonal spread of colistin resistance due to multiple mutational mechanisms in CPE [19]. Since the number of CPE strains has been increasing in Japan, it will be necessary to continually monitor the MIC of colistin in carbapenemase-producing Enterobacteriaceae strains[20].

Nine *mcr* genes in ESBL-producing Enterobacteriaceae and CRE strains were investigated in this study. The positive rate of *mcr* genes in all strains was 1.5%, which was similar to that in other previous reports from clinical samples (0.2-3.2%) [8, 21-24]. Therefore, in Japan, it seems that *mcr* genes have not become widespread in bacteria. One of the reasons for this is that colistin is currently used only in limited situations in Japan [25]. On the other hand, large amounts of colistin have been used in animals [26-28]. The first plasmid-mediated colistin resistance gene in Enterobacteriaceae, *mcr-1*, was detected in food-producing animals and humans in China, and it is likely that *mcr-1*-mediated colistin resistance originated in animals and subsequently spread to people [8]. In addition, the presence of plasmids containing *mcr* genes in *E. coli* from livestock animals has previously been reported in Japan. In particular, a high prevalence of *mcr-1* and *mcr-5* has been observed among strains isolated from diseased pigs [11]. Thus, we need to be wary of future spreading of *mcr* gene-positive strains in humans.

The *mcr-1* and *mcr-9* are distributed worldwide, *mcr-4, mcr-2* and *mcr-8* are slightly distributed, and other *mcr* are limited[29]. In this study, we identified three *mcr-9* positive *Enterobacter cloacae* complex. Although the *Enterobacter cloacae* complex coharboring carbapenemase gene and *mcr-9* has previously reported in Japan[30], carbapenemase genes were not detected in the three *mcr-9* positive stains. We also identified one *mcr-5* positive *E. coli*, but there were no reports of *mcr-5* positive *E. coli* infecting humans in Japan. The patient infected with *mcr-5* gene-positive strain had recurrent urinary tract infection, but the patient had never been treated with colistin. The acquisition of *mcr* gene-positive strains in humans has been reported to be transmitted from livestock, but the patient had no history of contact with animals. There was a possibility that *the mcr-5*-positive strain resulted from meat consumption in this case because the number of *mcr-5* positive strains isolated in livestock was higher in Japan than in other countries [11]. In this case, there was no history of treatment with colistin, whereas the patient was previously treated with levofloxacin and piperacillin-tazobactam. Since a previous study reported that the risk factors for *mcr-*positive Enterobacteriaceae were immunosuppression and history of antibiotic use, particularly carbapenems and fluoroquinolones [31], past use of levofloxacin may have been a risk factor for acquiring the *mcr-5* positive strain in this patient.

Whole-genome sequencing analysis revealed that the *mcr-5* positive strain was ST1642 and carried two distinct plasmids, the ESBL gene-carrying pN-ES-6-1 and *mcr-5*-carrying pN-ES-6-2. Although one study reported three *mcr-1-*harboring ESBL-producing *E. coli* ST1642 strains from bovine fecal samples[32], the *mcr-5*.*1*-harboring ESBL-producing *E. coli* ST1642 has not been reported previously. We found transposons of TnShfr1 (Tn3-family) in the *mcr-5* carrying *E. coli*. The *mcr-5*.*1* gene is reportedly located on the Tn3-familiy transposon of the *Salmonella enterica* Paratyphi B and *Cupriavidus gilardii* [33]. Therfore, there is a possibility that the *E. coli* strain received plasmid from these bacteria. In this study, the *mcr* genes-positive strain exhibited low MIC for colistin, which was not interpreted as resistant according to EUCAST. The results is the same as the previous reports [11, 34]. These results indicate that *mcr-5* and *mcr-9* may silently spread among Enterobacteriaceae, such as the stealth phenotype CPE [35].

This study has some limitations. We investigated the prevalence of colistin-resistant and plasmid-mediated colistin resistance genes in ESBL-producing and CRE strains. Since *mcr*-positive strains were also detected in non-ESBL-producing and non-CRE strains [36], we need to perform surveillance in all Enterobacteriaceae including drug-sensitive strains. In addition, we did not investigate intrinsically resistance to colistin.

In conclusion, we revealed that the rates of colistin-resistance and *mcr*-positive strains are not high in Japan. Since the MIC for colistin was low in the *mcr-*5 or *mcr-9* gene-positive strain, continuous monitoring of plasmid-mediated *mcr* genes in Enterobacteriaceae is necessary.

## Notes

### Author contributions

NK, NA, KS, KK, YM, and KY contributed to study design and data interpretation. K I, YY, HM, MK, and KO contributed to study design and collection of isolates and data. N. K. provided expert advice, critically reviewed the manuscript, including for aspects related to the English language, and contributed to its content. All authors reviewed and approved the final version of the manuscript.

### Disclaimer

The funders had no role in the design of the study, the collection or analysis of data, or the decision to publish.

### Financial support

This work was supported by the Japanese Society of Laboratory Medicine Fund for the Promotion of Scientific and the Health and Labour Sciences Research Grants from the Ministry of Health, Labour and Welfare, Japan (H28-Shinkou-Ippan-003).

### Potential conflicts of interest

All authors: No reported conflicts of interest.

## Notes

### Competing Interest Statement

The authors have declared no competing interest.

