## Supplemental Tables for "Surveillance of colistin-resistance and *mcr genes in* multi-drug resistant Enterobacteriaceae"

**Supplementary data**

**Table S1.** Characteristics of colistin resistant strains

| Characteristics | Resistance of colistin |
| --- | --- |
| Region where the strain was isolated |  |
| Western Japan (A hospital) | 8/90 (8.9%) |
| Central Japan (B and C hospitals) | 4/65 (6.2%) |
| Eastern Japan (D and E hospitals) | 9/118 (7.6%) |
| Sites of infection |  |
| Urinary tract | 6/120 (5.0%) |
| Intra-abdominal | 3/41 (7.3%) |
| Respiratory tract | 8/55 (14.5%) |
| Blood | 1/23 (4.3%) |
| Stool | 2/20 (10.0%) |
| Others | 1/14 (7.1％) |

**Table S2.** Antibiotic susceptibility testing of *mcr* gene positive trains

| Antibiotics | *mcr-5*  *E. coli*  ESBL | *mcr-9*  *Ent. cloacae*  CRE | *mcr-9*  *Ent. cloacae*  CRE | *mcr-9*  *Ent. cloacae*  CRE |
| --- | --- | --- | --- | --- |
| Piperacillin | >64 | ≤2 | 16 | 8 |
| Ceftazidime | 2 | ≤1 | 8 | 16 |
| Ceftriaxone | ≤0.5 | ≤0.5 | 2 | 2 |
| Cefpodoxime | >4 | ≤4 | >4 | >4 |
| Cefepime | ≤0.5 | ≤1 | ≤1 | ≤1 |
| Cefmetazole | 32 | >32 | >32 | >32 |
| Aztreonam | 4 | 2 | 8 | >32 |
| Piperacillin-tazobactam | 16 | ≤2 | 16 | 16 |
| Ampicillin-sulbactam | >16 | ≤2 | 16 | 16 |
| Imipenem | ≤0.5 | 2 | 2 | 2 |
| Meropenem | ≤0.5 | ≤0.5 | ≤0.5 | ≤0.5 |
| Gentamicin | >8 | ≤0.5 | ≤0.5 | ≤0.5 |
| Amikacin | ≤4 | ≤2 | ≤2 | ≤2 |
| Minocycline | >8 | 2 | 2 | 4 |
| Ciprofloxacin | >4 | ≤0.5 | 2 | ≤0.5 |
| Levofloxacin | >4 | ≤1 | 2 | ≤1 |
| Sulfamethoxazole-trimethoprim | >4 | ≤1 | ≤1 | ≤1 |
| Colistin | 1 | ≤0.5 | ≤0.5 | ≤0.5 |
